# Macrolinguistic function in temporal lobe epilepsy: a reinterpretation of circumstantiality

**DOI:** 10.1101/2022.08.22.22279104

**Authors:** Fiore D’Aprano, Charles B. Malpas, Stefanie E. Roberts, Michael M. Saling

## Abstract

Individuals with temporal lobe epilepsy (TLE) often have an impairment of conversational language manifesting as verbosity and attributable to disruption of cognitive-linguistic networks. The micro- and macrolinguistic underpinnings of this disturbance, and the role of epilepsy and cognitive variables, are yet to be explored. We examined the elicited language output of 16 individuals with TLE and 14 healthy controls under separate monologic discourse tasks: a structured and constrained context, elicited by description of the ‘Cookie Theft’ picture, and an unstructured, unconstrained context, elicited by description of a ‘Typical Day’. We hypothesised that language output in the unstructured context would be characterised by verbosity to a greater extent than language elicited in a structured context. Following transcription and coding, detailed multi-level discourse analysis suggested that a constrained context gives rise to microlinguistic disturbances in individuals with TLE, reducing fluency, with more pauses and fillers. Under an unconstrained context, as anticipated, classical aspects of verbosity emerge in those with TLE, manifesting as longer speaking time, a longer duration of pauses, and a higher proportion of repetitive or redundant statements. Macrolinguistic elements such as coherence and informativeness are widely impacted, particularly disturbing language formulation. Correlations suggest that microlinguistic disturbances are closely linked with the immediate impact of seizures on cognitive-linguistic function, while macrolinguistic disturbances are more broadly impacted by disorder severity and word retrieval deficits. These findings suggest that different psycholinguistic impairments emerge as a function of differing linguistic challenges imposed by constrained and unconstrained conversational contexts. We conclude that these patterns reflect a dynamic linguistic system taking shape under specific contextual conditions.

**Highlights:**

- A multi-centre multi-level discourse analysis was carried out to examine language in temporal lobe epilepsy (TLE).
- Distinct linguistic impairments reflect task demands which differentially affect micro- and macrolinguistic features.
- In constrained conditions, individuals with TLE exhibit microlinguistic disturbances mainly affecting their fluency and output clarity.
- In unconstrained conditions, they are verbose and have disturbed coherence where they produce more content that is repetitive or redundant.

## 1. Introduction

Individuals with temporal lobe epilepsy (TLE) exhibit characteristically verbose language [1]. This pedantic, repetitive, highly-detailed, and peripheral style is termed ‘circumstantiality’ and was initially considered a personality feature of TLE [1,2]. It is now better conceptualised as a non-lateralised neurocognitive phenomenon attributable to subtle interictal disruptions of linguistic function. While language impairments in TLE are commonly identified clinically, proffered as a cognitive complaint, or detected on neuropsychological assessment at a single-word level [3,4], there is limited understanding of how these characteristic impairments relate to dysfunctional discourse.

Circumstantiality in TLE has been proposed to serve as a compensatory mechanism to overcome instances of word-finding difficulties [5,6]. This is disputed by Field and colleagues [7] who posit that lexical-processing deficits in left TLE do not account for circumstantiality, suggesting that micro- and macrolinguistic processes are dissociable, being those that relate to lexical-syntactic and suprasentential processes, respectively. Discourse processing deficits in narrative and conversational contexts are also found in right TLE, and are dissociable from lexical-syntactic impairments [8]. Similar patterns of macrolinguistic dysfunction are present following right hemisphere damage, particularly accompanying anterior lesions [9]. While their microlinguistic function appears intact, impairments emerge in coherence, producing more tangential output [9]. Right frontal regions, as components of complex networks, may therefore play a role in organising information in narrative discourse [9]. Disrupted projections from temporal to frontal regions in TLE [10] might engender similar patterns as those observed by Marini [9].

Fluency, cohesion, and coherence have been largely overlooked by TLE researchers to date. In this context, fluency, a microlinguistic element, relates to instrasentential or message-level transitions, and describes the rate of language production, rather than psychometrically-evaluated verbal fluency on tasks of orthographic lexical retrieval or semantic fluency [11]. Individuals with left TLE tend to pause longer than those with right TLE [12], and regardless of laterality, they use more noncommunicative fillers, abandon trains of thought, and produce more repetitions than controls [13]. Despite more frequent fluency disruptions, they are comparable with healthy controls in their *rate* of output, that is, words per minute [7,12]. Cohesion and coherence are macrolinguistic elements. Cohesion describes relatedness of meaning across sentences by making explicit and unambiguous connections to previously introduced content. Cohesive devices include conjunctions and personal referents; for example, *Sam was hungry. <u>He</u> made dinner* [14]. Cohesion relies on simultaneously monitoring output and the listener’s understanding which depends upon working memory [15] and lexical retrieval [16]. Global coherence relates to discourse organisation above the sentence-level, that is, how content is connected thematically to serve a particular goal or plan [17].

Demands on language production are thought to be influenced by type of elicitation (Fergadiotis & Wright, 2011; Stark et al., 2019). Structured elicitation tasks, such as describing a cartoon, provide a clear discourse topic and adequate linguistic scaffolding to facilitate discourse planning and limit disturbances. In a structured narrative production task, participants with TLE were not verbose when describing a six-frame cartoon, and instead were comparable to healthy controls in the number of words and output duration, but were less fluent overall [13]. There was no evidence to suggest that these disturbances to discourse related to laterality of seizure onset [13]. Using the structured ‘Cookie Theft’ task [18] Hoeppner and colleagues [19] reported verbosity in four of nine participants with left-localised focal impaired awareness seizures (FIAS). They produced more words, dysfluencies, and non-essential details. Rather than verbosity being a broad diagnostic feature in TLE, structure might facilitate discourse processing in some individuals. While a subset were verbose in this structured task, verbosity might be more widely distributed in an unstructured context, such as eliciting spontaneous output in response to an open-ended question, which likely disturbs macrolinguistic components regarding planning and coherence [20]. These processes rely on disparate aspects of cognition and given network dysfunction in TLE, discourse disturbances are unlikely lateralised [16]. Unstructured contexts are more representative of communicative function [21] and are akin to the spontaneous language produced clinically where unusual language features in TLE are anecdotally reported.

The examination of naturalistic output that we will describe here is novel and addresses the paucity of detailed, systematic, and appropriate investigation of language in this population. This study aimed to characterise language impairments in TLE by examining discourse under structured (Cookie Theft) or unstructured (Typical Day) conditions. To date, spontaneous output in TLE has been largely examined with aphasia batteries which are appropriate to detect focal impairments following stroke, but are not, by design, sensitive to the subtle language changes that characterise TLE [4]. Given that microlinguistic processes are language-specific, and underpinned by subtle interictal disturbances [22], we hypothesised that individuals with TLE would have microlinguistic disturbances in fluency, marked by more frequent hesitations and false starts, irrespective of contextual demands. On the other hand, given the reliance of macrolinguistic processes on broader aspects of cognition including planning and working memory [16,23], we anticipated that macrolinguistic disturbances would emerge when there were fewer constraints on output and therefore greater demands on high-level language systems: individuals with TLE would be comparatively verbose, using more words and speaking for a longer duration, and would additionally exhibit disturbances to cohesion and coherence.

## 2. Material and Methods

### 2.1 Participants

This study included 30 participants, 16 with focal unilateral TLE (comprising nine mesial), and 14 age-sex- and education-matched healthy controls. These individuals experience seizures of temporal lobe origin, typically focal aware seizures (FAS) or FIAS and are patients of either The Royal Melbourne Hospital or the Alfred Hospital in Melbourne, Australia. Diagnoses are made as part of a comprehensive team in accordance with International League Against Epilepsy criteria [25]. Consensus on the basis of seizure semiology, video electroencephalography, magnetic resonance imaging, positron emission tomography, and inter-ictal single-photon emission computer tomography provided unambiguous diagnoses with a localised seizure focus, 11 of which had left hemisphere involvement.

Inclusion criteria were: English as a first language; a diagnosis of drug-resistant TLE at the time of recruitment [24]; no prior neurosurgical resection; full scale IQ >70; no reported history of substance-related and addictive disorders, no formally diagnosed psychiatric disorders, and not currently experiencing a major psychiatric episode (e.g., psychosis). None of these individuals were receiving additional treatments for the control of seizures (e.g., vagal nerve stimulation) and none had a history of developmental language disorder or other neurological condition (e.g., stroke). By assessment, three individuals with TLE reported no seizures in the preceding 12 months on their current anti-seizure medication (ASM) regimen [24]. Analyses were performed both with and without these drug-responsive participants to determine whether their inclusion in the sample was a robust choice. We found no difference in key outcome measures and these individuals were subsequently retained in the sample. Their reduced epilepsy burden at that point in time is reflected in the 13-point Seizure Frequency Rating [26]; considering seizure frequency, type, and ASM usage. Healthy controls comprised family members or partners of TLE participants, and where necessary were recruited from the community via convenience sampling to ensure the groups were demographically comparable by appropriately age-, education-, and sex-matching to those with TLE.

This multi-site study received ethical approval from the Melbourne Health Human Research Ethics Committee in accordance with ethical standards of the 1964 Declaration of Helsinki. All participants provided written informed consent.

### 2.2 Neuropsychological Assessment and Discourse Elicitation

This study involved neuropsychological and language assessment (Table S2), conducted by a single registered psychologist. Given the COVID-19 lockdown conditions in Melbourne, Australia at the time of collection, these assessments were predominantly completed via telehealth (Table 1). Lexical retrieval was assessed using the Boston Naming Test (BNT) [27], the Controlled Oral Word Association Test (COWAT) [28], the Auditory Naming Test (ANT) [29] with minor modifications to suit the Australian lexicon, and the Verb Generation Task (VGT) developed to examine verb retrieval (Appendix S1).

**Table 1.** Sample characteristics for TLE group and healthy controls

| Variable | Control<br>(*n* = 14) |  | TLE<br>(*n* = 16) |  | *p* | Effect size |
| --- | --- | --- | --- | --- | --- | --- |
|  | Median (*Q1*, *Q3*) | Range | Median (*Q1*, *Q3*) | Range |  |  |
| Age [years] | 51.50 (26.00, 55.50) | 50 | 43.50 (32.50, 55.50) | 44 | 0.92 | 0.03 |
| Education [years] | 15.50 (12.00, 18.00) | 9 | 14.00 (11.75, 17.00) | 15 | 0.45 | 0.17 |
| Estimated IQ<sup>a</sup> | 107.75 (97.63, 111.88) | 41 | 103.75 (95.88, 107.00) | 50.50 | 0.20 | 0.28 |
| Age at Diagnosis [years] | N/A | N/A | 26.50 (21.00, 46.25) | 50 |  |  |
| Epilepsy duration [years] | N/A | N/A | 7.50 (3.50, 16.25) | 41.50 |  |  |
| Seizure Burden Rating | N/A | N/A | 6.50 (2.00, 7.00) | 8 |  |  |
| Last seizure [weeks] | N/A | N/A | 10.00 (4.00, 34.00) | 75 |  |  |
| Current ASMs | N/A | N/A | 2.00 (1.00, 2.00) | 3 |  |  |
| Laterality, Left [*n*, %] | N/A | N/A | 11 (69) |  |  |  |
| Mesial focus [*n*, %]<sup>^</sup> | N/A | N/A | 9 (56) |  |  |  |
| Sex, Female [*n*, %] | 7 (50) |  | 7 (44) |  | 0.73 | 0.06 |
| Handedness, Right [*n*, %] | 12 (86) |  | 13 (81) |  | 0.74 | 0.06 |
| Telehealth [*n*, %] | 14 (100) |  | 15 (94) |  | 0.34 | 0.17 |

*Note.* TLE = Temporal Lobe Epilepsy; ASM = Anti-seizure Medication. For continuous variables, group differences were computed using Mann-Whitney U test, effect size reported is the rank biserial correlation coefficient. <sup>a</sup>Suggests that data does not violate assumptions of normality on Shapiro-Wilk. For discrete variables, percentages of each group reported, group differences computed using X<sup>2</sup> test of independence, effect size reported is phi-coefficient. Estimated IQ reflects a composite between TOPF scaled score and WASI-II FSIQ-2 score. Seizure frequency calculated on 13-point rating scale (0-12) considering ASM usage, frequency, and type of seizure (So et al., 1997). <sup>^</sup>Of these individuals 5 had a diagnosis of left TLE. Non-mesial cases comprise 4 neocortical (3 left, 1 right), and 3 non-lesional.

Discourse-level language was examined via two forms of elicitation: one being unstructured and unconstrained (Typical Day) by prompting participants “tell me about a typical day in your life”; and the other being narrative-based, structured, and highly constrained in its content (Cookie Theft of the Boston Diagnostic Aphasia Examination; [18]). The cartoon image was presented on an A4 sheet of paper at the bedside, and via screen sharing for telehealth assessments and remained visible throughout their description to minimise any memory contribution. Participants were encouraged to take enough time to understand the cartoon and were prompted to “tell me everything you see happening in this scene”. There was no time limit imposed for either task and the order of presentation was identical for all participants. The researcher minimised verbal and non-verbal participation and participants were required to make a definitive statement to conclude the task e.g., “I’m finished”. Participants were prompted “is there anything else?” where a particularly brief, simplified description reasonably indicated a misunderstanding of task requirements, or where greater than five seconds of silence had elapsed without self-disclosing their completion.

### 2.3 Recording and transcription

Language samples were audio recorded for subsequent transcription and analyses. A Yeti microphone and Audacity® software was used at the bedside, while telehealth output was recorded from the Zoom session [30]. All audio files were then manually transcribed verbatim and segmented by a single researcher within four weeks of the file being obtained. Statement segmentation aligns with methodology applied by Stein and Glenn [31] and Trabasso and van den Broek [32], where a single statement refers to a predicate and its corresponding arguments. This promotes consistent proposition-based extraction of content [33], rather than by communicative unit (C-unit). Pause lengths were manually extracted from Audacity^®^ software. Sample lengths refer to the total number of completed words, excluding words filling pauses such as “um” “uh” [34,35].

### 2.4 Discourse variables and coding practices

Based on models of discourse production and discourse examination in clinical populations, select discourse variables were used as the basis for the discourse analysis. These relate to key linguistic components at both the micro- and macrolinguistic level, being lexical-syntactic and suprasentential, respectively. A description of all nodes coding is presented in Table S3. Transcript coding was completed using NVivo 12 software by a single researcher who was blind to participant characteristics, other than data acquisition date. Each transcript was coded twice by the same reviewer, with an intra-rater agreement of 94% across coding occasions. Where discrepancies were identified, the researcher re-considered the criteria to produce a final decision. Any ambiguities in the description of criteria for coding to nodes were clarified.

### 2.5 Coding agreement

A second expert researcher was assigned to blindly code a random subset of transcripts in NVivo. In line with other similar discourse analyses, this was determined to be 12.5% [36] and included five transcripts of each task. Inter-rater agreement was determined on a point-by-point basis for each node to allocate as well as appropriate statement segmentation. The second researcher had access to the complete, disambiguated code-book for this process (Table S3). For nodes, percentage of agreement was 90%, while for the segmentation of statements, that is, determining each predicate and its corresponding arguments, agreement was 92%, surpassing the minimal accepted requirement level of 80% [37]. Coding discrepancies were discussed among the reviewers and resolved via consensus. Once again, any ambiguities in node descriptions were resolved and their coding was updated (Table S3).

### 2.6 Statistical Analyses

Jamovi software [38] was used for all analyses. For sample characteristics, neuropsychological measures, and discourse variables group-specific measures of central tendency and group-differences were computed. Many of the data were skewed and to be conservative, non-parametric tests (Mann-Whitney U test) have been applied across all analyses for the purpose of uniformity. Data that did not violate assumptions of normality are indicated in table notes. Contingency tables (X^2^ test of independence) were used for categorical variables. The rank biserial correlation (RBC) was used as a non-parametric estimate of effect size, reflected as small 0.1 < 0.3 < 0.5 large. Micro- and macrolinguistic discourse variables that significantly distinguished individuals with TLE from controls were then subject to correlation analyses, to examine their relationship to clinical and cognitive characteristics, and are reported as Spearman’s rank correlation coefficients. To ensure that the analyses were not sensitive to an artifact such as group, we ran the analyses with left and right TLE separately and then together. These analyses suggested no difference in discourse outcomes between left and right TLE or relative to healthy controls when individuals with TLE were considered as separate left and right groups or a single group. Consistent with the notion that high-level language functions are not lateralised, all individuals with TLE were subsequently treated as a single group. This methodological point is further addressed in the discussion. To account for Type I error, a false detection rate (FDR) of 0.05 was applied to primary analyses [39].

## 3. Results

### 3.1 Sample characteristics

Individuals with TLE and healthy controls were comparable across most demographic characteristics and many aspects of neuropsychological function (Table 1 and Table S2). Those with TLE reported higher rates of depressive symptomatology than healthy controls. A lexical retrieval deficit broadly emerged in TLE, and they reported more frequent and distressing word finding difficulties than controls. Rather than total raw or scaled scores for the number of correct items, *delays* in word retrieval for BNT and ANT appeared to be more sensitive to failures of lexical retrieval—increased mean response time latencies and increased tip-of-the-tongue (TOT) states, i.e., responding >2000ms post-stimulus *or* requiring phonemic prompting. Individuals with TLE also demonstrated longer latencies on VGT. Relative to controls, individuals with TLE also produced fewer words within a semantic category. See Table S2 (Supplementary Material).

### 3.2 Cookie Theft: Constrained Output

The groups were comparable in most aspects regarding output volume and coherence, including sample length, duration of output, duration excluding pauses, total number of statements, total core units, proportion of non-progression to novel statements, and syntactic simplicity. There was the strongest statistical evidence for group differences in fluency (see Table 2), as these relationships held when FDR correction was applied. This included individuals with TLE producing significantly more fluency disruptions, which include more false starts, clarity disruptors, and hesitations such as pausing more frequently at non-grammatical junctures when compared to healthy controls. There was weaker evidence for disturbed cohesion, pause duration, and production rate which did not survive correction. Individuals with TLE produced more elements that disrupted cohesion, including more ‘other’ referents, which are incomplete, ambiguous, or missing. There was some indication of a longer duration of pauses, and a slower production rate than healthy controls. The relative ranking of effect sizes are presented in Figure 1. Examples of this output are provided in Supplementary Material, Table S4.

**Table 2.**
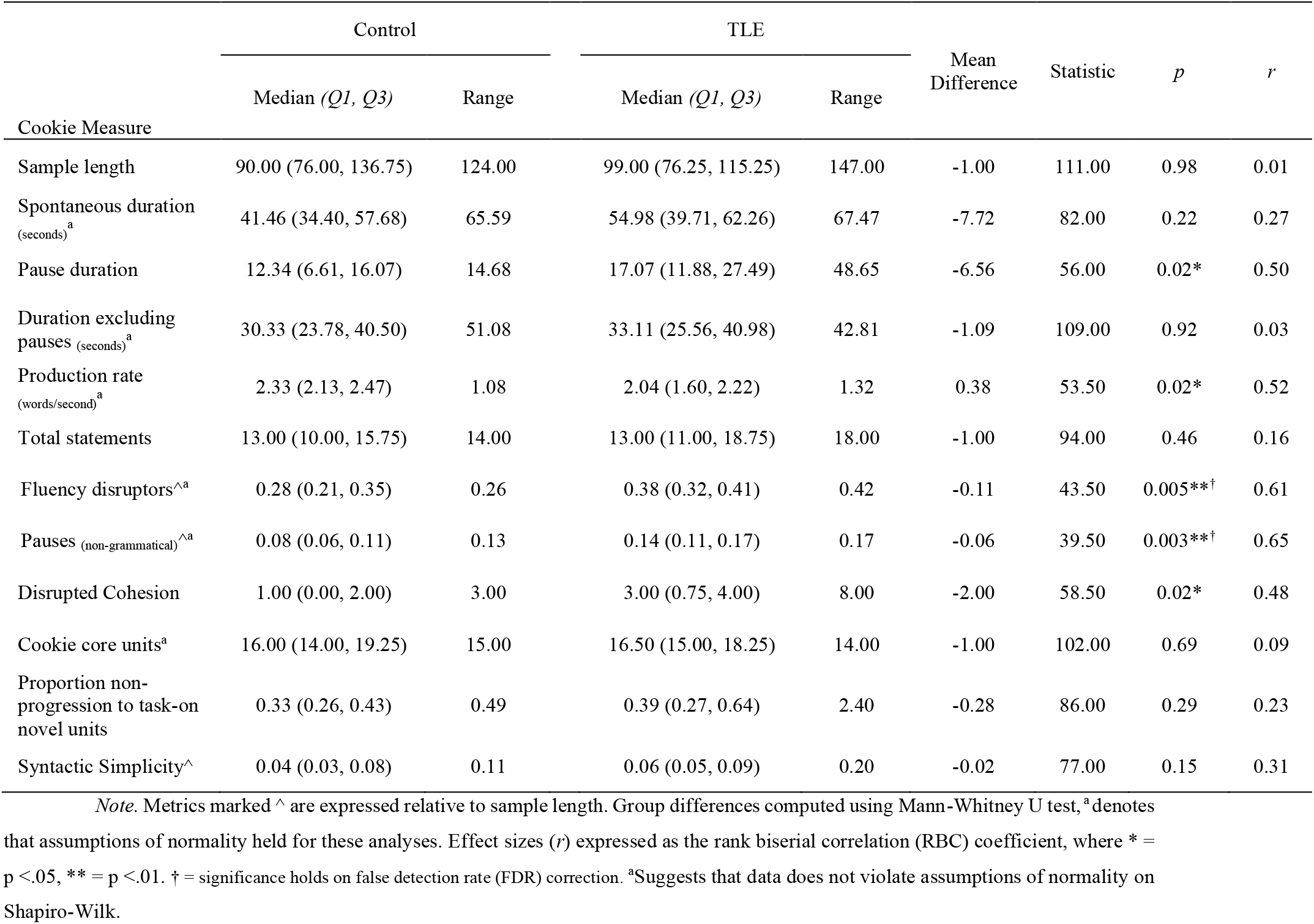
Group differences in discourse features in constrained output: Cookie Theft task

**Figure 1.**
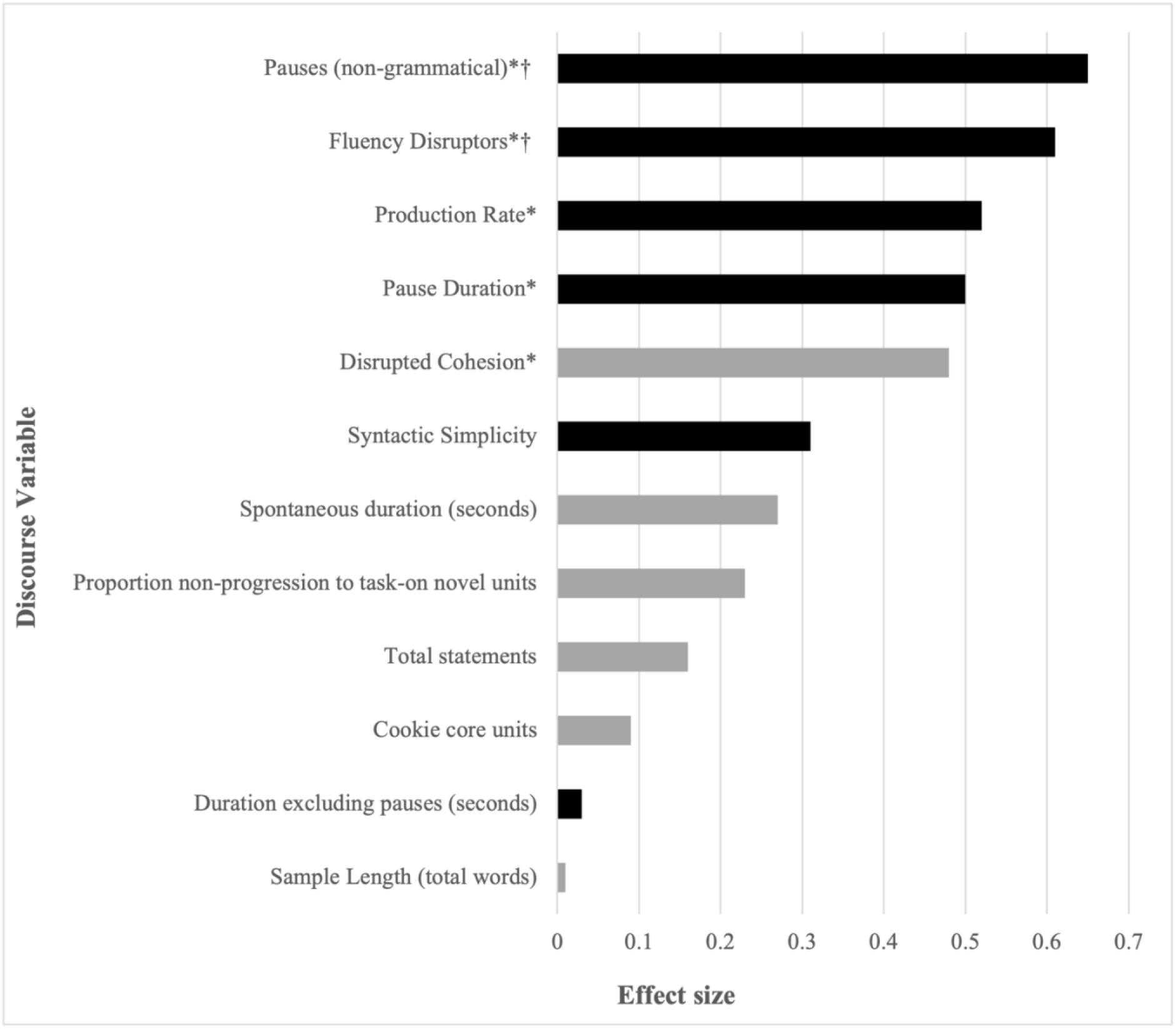
Mean differences on discourse variables between TLE and Healthy Controls for Cookie Theft task, represented as absolute value effect size (*r*, small 0.1 < 0.3 < 0.5 large), * = significant difference, † = difference holds on FDR correction. Microlinguistic features are represented in black, while macrolinguistic features are grey.

### 3.3 Typical Day: Unconstrained Output

Group differences emerged in coherence, informativeness, and circumstantiality of output (see Table 3). The relative ranking of effect sizes can be visualised in Figure 2. Examples are provided in Supplementary Material, Table S5. There was the strongest statistical evidence for group differences in spontaneous duration, pause duration, and proportion of non-progression to novel units, which remained significant on FDR correction. Compared to healthy controls, individuals with TLE spoke for longer and had a longer duration of pauses. With regards to informativeness, those with TLE had a higher proportion of non-progression statements to task-on novel statements than healthy controls, that is, they produced more statements that did not progress content than statements that did. There was weaker evidence for a longer sample length in TLE, more frequent pauses at non-grammatical junctures, and a longer duration excluding pauses, suggesting that the longer duration of pauses in TLE did not account for the difference in spontaneous duration. While those with TLE produced more total statements, this difference was not significant.

**Table 3.** Group differences in discourse features in unconstrained output: Typical Day task

| Cookie Measure | Control |  | TLE |  | Mean Difference | Statistic | *p* | *r* |
| --- | --- | --- | --- | --- | --- | --- | --- | --- |
|  | Median (Q1, Q3) | Range | Median (Q1, Q3) | Range |  |  |  |  |
| Sample length | 134.50 (92.00, 203.25) | 298.00 | 188.00 (143.00, 453.00) | 546.00 | -73.50 | 59.00 | 0.04* | 0.44 |
| Spontaneous duration<sup>a</sup><br>(seconds) | 57.43 (43.65, 85.15) | 89.13 | 89.26 (64.83, 191.66) | 211.04 | -42.38 | 50.00 | 0.01*† | 0.52 |
| Pause duration<sup>a</sup> | 14.64 (10.57, 18.96) | 26.19 | 28.23 (19.06, 53.53) | 54.76 | -13.64 | 34.00 | 0.001**† | 0.68 |
| Duration excluding pauses<sup>a</sup><br>(seconds) | 40.74 (31.70, 62.30) | 84.52 | 62.22 (45.76, 134.73) | 158.67 | -29.10 | 54.00 | 0.03* | 0.49 |
| Production rate<sup>a</sup><br>(words/second) | 2.33 (2.04, 2.68) | 1.24 | 2.30 (2.15, 2.50) | 1.28 | 0.13 | 93.50 | 0.63 | 0.11 |
| Total statements | 25.00 (17.25, 31.50) | 50.00 | 29.00 (24.50, 70.0) | 29.00 | -11.00 | 65.50 | 0.09 | 0.38 |
| Fluency disruptors<sup>^a</sup> | 0.34 (0.27, 0.36) | 0.41 | 0.36 (0.28, 0.42) | 0.36 | -0.05 | 79.00 | 0.27 | 0.25 |
| Pauses<sup>(non-grammatical)^a</sup> | 0.10 (0.08, 0.12) | 0.16 | 0.14 (0.11, 0.16) | 0.14 | -0.03 | 51.00 | 0.02* | 0.51 |
| Disrupted Cohesion | 1.00 (0.00, 6.75) | 13.00 | 4.00 (3.00, 6.50) | 15.00 | -2.00 | 68.50 | 0.11 | 0.35 |
| Proportion non-progression to task-on novel units | 0.39 (0.24, 0.51) | 0.95 | 0.78 (0.51, 0.91) | 3.17 | -0.35 | 45.00 | 0.009**† | 0.57 |
| Syntactic Simplicity<sup>^a</sup> | 0.11 (0.03, 0.13) | 0.20 | 0.10 (0.06, 0.11) | 0.17 | 0.01 | 98.50 | 0.79 | 0.06 |

*Note.* Metrics marked ^ are expressed relative to sample length. Group differences computed using Mann-Whitney U test, <sup>a</sup> denotes that assumptions of normality held for these analyses. Effect sizes (*r*) expressed as the rank biserial correlation (RBC) coefficient, where \* = *p* < .05, \*\* = *p* < .01. † = significance holds on false detection rate (FDR) correction. <sup>a</sup>Suggests that data does not violate assumptions of normality on Shapiro-Wilk.

**Figure 2.**
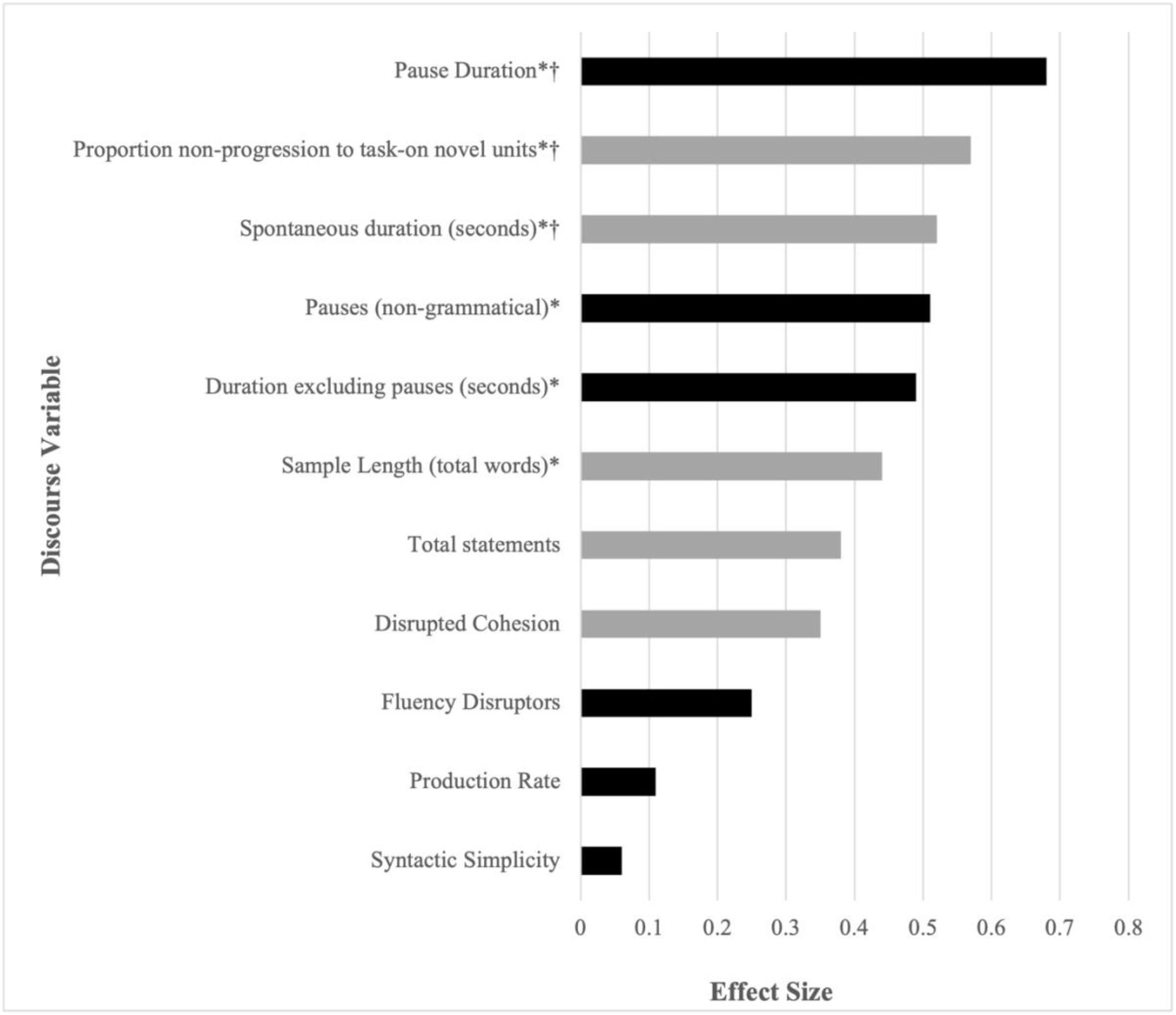
Mean differences on discourse variables between TLE and Healthy Controls for Typical Day task, represented as absolute value effect size (r, small 0.1 < 0.3 < 0.5 large), * = significant difference, † = difference holds on FDR correction. Microlinguistic features are represented in black, while macrolinguistic features are grey.

### 3.4 Correlations

The relationships between these discourse variables surviving FDR correction and demographic, epilepsy-specific, and cognitive variables are reported in Table 4.

**Table 4.** Correlations between key discourse variables, demographic, clinical, and cognitive characteristics

| Variables of interest | Cookie Theft |  | Typical Day |  |  |
| --- | --- | --- | --- | --- | --- |
|  | Fluency disruption | Pauses<br>(non-grammatical) | Spontaneous<br>Duration | Pause<br>Duration | Non-progression to<br>novel units |
| Age at assessment | -0.01 | -0.08 | -0.05 | -0.13 | 0.14 |
| Years of Education | -0.15 | 0.08 | -0.37 | -0.23 | -0.33 |
| Disease duration | 0.50**† | 0.47** | 0.43* | 0.55**† | 0.57**† |
| Time since last seizure | 0.58***† | 0.56**† | 0.41* | 0.53**† | 0.48**† |
| Seizure frequency rating | 0.40* | 0.47** | 0.46*† | 0.57**† | 0.46*† |
| Number of ASMs | 0.44* | 0.48** | 0.48*† | 0.58**† | 0.63**† |
| IQ composite | -0.25 | -0.00 | -0.43* | -0.42** | -0.25 |
| Victoria Stroop Dots | -0.15 | -0.09 | -0.11 | -0.03 | 0.35 |
| WAIS-IV Digits Backwards | 0.19 | 0.19 | -0.37* | -0.21 | -0.16 |
| Animals | -0.22 | -0.20 | -0.20 | -0.36 | -0.25 |
| OLR | -0.30 | -0.09 | -0.24 | -0.31 | -0.01 |
| Lexical retrieval TOT | 0.40* | 0.33 | 0.40* | 0.44* | 0.50**† |
| Lexical retrieval latency | 0.20 | 0.21 | 0.38* | 0.41* | 0.32 |
| HADS depression | 0.13 | 0.27 | 0.19 | 0.31 | 0.38* |

*Note.* ASM = Anti-seizure Medication; IQ = Intelligence Quotient; WAIS-IV = Wechsler Adult Intelligence Scale, Fourth Edition; WMS-IV = Wechsler Memory Scale, Fourth Edition; OLR (COWAT) = Orthographic Lexical Retrieval (Controlled Oral Word Association Test); TOT = tip of the tongue state; HADS = Hospital Anxiety and Depression Scale. Reported as Spearman rank correlation coefficients. \*p < .05, \*\* p < .01, \*\*\* p < .001. † = hold on FDR correction.

#### 3.4.1 Cookie Theft Correlations

Microlinguistic components distinguished individuals with TLE from healthy controls and relate closely to seizure characteristics. Fluency disruptions correlated significantly with epilepsy variables including disease duration and time since last seizure. The number of non-grammatical pauses also significantly correlated with the time since their last seizure.

#### 3.4.2 Typical Day Correlations

All three variables of interest, including micro- and macrolinguistic elements, correlate strongly with epilepsy-specific variables relating to seizure frequency and the number of ASMs. Pause duration and the proportion of non-progression to novel units are also related to disease duration and the time since last seizure. The increased proportion of non-progression units correlates with more TOT states.

## 4. Discussion

Our findings suggest that individuals with TLE are not more impaired in one communicative context than another, but rather distinct linguistic impairments emerge and are hypothesised to reflect task demands. One task is highly constrained, structured, visually prompted, and contains a focused and finite set of elements to communicate, while the other is highly unconstrained, unstructured, and therefore particularly vulnerable to the effects of verbosity.

Under conditions of constrained output, there is a specific focus on providing a complete description of the visual stimulus. To produce a narrative, individuals are limited in their lexical choices and direction of output to meet task demands, and therefore have less agency and less variability in their output. According to our findings, the restricted scope of discourse poses a linguistic challenge to individuals with TLE, who might otherwise change direction when faced with a lexical retrieval deficit. In this context their fluency is predominantly impacted, cohesion to a lesser extent, and no overall pattern of circumstantiality. The strongest evidence was for disturbed fluency, including more frequent false starts, clarity disruptors and non-grammatical hesitations than in healthy controls, reflecting a disturbance overall output quality. In this context, dysfluencies plausibly represent compensations for lexical retrieval deficits. These findings are consistent with those of both Hoeppner and colleagues [19] who found greater dysfluencies on this Cookie Theft task across *all* individuals with FIAS, and in Bell and colleagues [13] on a narrative task. While there was some suggestion in our findings that production rate was slower in TLE, at this stage we do not have robust evidence to suggest any difference in disturbed output rate relative to healthy controls, consistent with previous findings [7,12]. While Hoeppner and colleagues [19] reported a verbose subset of participants with left-localised FIAS, Bell and colleagues [13] suggested that individuals with TLE were similar to healthy controls in speaking time and total number of words, and were therefore not verbose. These are consistent with the present finding that there was no evidence of circumstantiality among individuals with TLE in this context.

The overall hypothesis was that verbosity, and ultimately macrolinguistic dysfunction, were more likely to be featured when output was spontaneous and unconstrained, and is supported by our findings. Individuals with TLE produced more microlinguistic errors in total, although these errors are proportional to those in healthy controls when considering sample length. While unstructured contexts have higher planning demands [12], in the current unconstrained context, content is familiar and therefore individuals are likely to have a more specific and refined lexicon for their personal lives, within a known semantic space. Presumably this familiarity limits the impact of lexical retrieval deficits on fluency. By virtue of saying more, their output does not seem as disrupted in its fluency as we might anticipate. With the freedom of spontaneous discourse, a different set of linguistic challenges emerge where macrolinguistic deficits prevail among those with TLE, impacting suprasentential mechanisms. According to our findings, they tend to deviate more from the topic of conversation; producing longer output, pausing for longer duration, and being less concise and informative in the content they communicate, that is, they say more about less. Producing more repetitive and redundant units that do not contribute to content progression disrupts coherence and represents a communicative issue. At a non-linguistic level, it has also been hypothesised that verbosity in TLE might have a social desirability component. In attempting to maintain social contact and be understood they become particularly repetitive and loquacious [22]. A constrained, impersonal task such as the Cookie Theft might not elicit verbose output given a weaker social-motivation component, potentially because the task is straight forward, largely invariable in its overall portrayal, not reliant on self-representation, and not followed by questions about its content. In contrast, the unstructured questioning regarding a Typical Day is more likely to elicit this social need to maintain face via elaboration, clarification, and self-disclosures irrelevant to task goals.

Discourse production is cognitively demanding [40]. Importantly, microlinguistic processes are considered language-specific, and underpinned by subtle interictal disturbances [22]. The present findings regarding microlinguistic disturbances align closely with our understanding of cognitive and linguistic systems, where we see differences in aspects of function that are presumed to be directly modified and compromised by seizures generally. For example, seizure burden, ASM usage, and their impact on processing speed [41,42] have ramifications for disrupting fluency, particularly in relation to non-grammatical pauses and pause duration, which are more highly determined by fundamental neurolinguistic systems, and correlate significantly with disease duration and recent seizures. These disturbances are disproportionately observed among individuals with TLE, regardless of context. On the other hand, syntactic complexity is presumed and found to be comparable between groups as it is socio-linguistically determined [43] and therefore remote from the effects of seizure activity. Instead, macrolinguistic processes rely on broader aspects of cognition [16]. As a network disorder, these systems are vulnerable to disturbances in TLE with seizure focus and propagation into distal regions [23]. The increased demands on linguistic and non-linguistic cognitive systems involved in planning and organising spontaneous discourse potentially account for the observation that macrolinguistic impairments become more overt when task constraints decrease, but might not be sufficiently captured by the limited scope of psychometric measures. These macrolinguistic disturbances are more vulnerable to the broader syndromal effects specific to TLE, including neurotoxicity and the impact of seizure activity and subsequent widespread cortical dysfunction which confer a degree of compromise in high-level language systems. This is mirrored in significant correlations between spontaneous duration and proportion of non-progression units with seizure frequency and number of ASMs, as indicators of disorder severity, as well as increase in TOT states being correlated with proportionally higher-non-progression units.

The sample size is in keeping with similar studies and is appropriate for the scope of a discourse analysis which focuses on thoroughly characterising language use [44]. Participants were well-matched on demographic characteristics. Of note, each discourse type was elicited using a single task and it is difficult to discern whether something inherent in these tasks might have given rise to these findings. However, examining errors relative to sample length, rather than raw values, has adequately equated these tasks to make appropriate comparisons. Future research could consider including multiple tasks with similar demands to confirm these findings. This study is concerned with high-level linguistic function which is not represented in brain tissue in a modular or focal fashion. There are many senses in which right and left TE are cognitively and linguistically distinct. While more fundamental aspects of cognitive-linguistic function can be localised or lateralised, this is not the expectation for higher-order language, as a higher cortical function. When dealing with linguistic function there is no a priori reason for thinking about this as a lateralised function. Functions as complex as this are seldom as localised as lateralised as one might think, and instead relate to widely distributed networks. In light of this, right and left TLE participants have been considered as a single diagnostic group. While this might sound counterintuitive, there was no sense in which the groups were distinguishable. This approach was supported by our analyses which indicated that there were no differences in discourse outcomes when considered as separate groups or collectively, and ultimately that treating them as a single group was a robust choice. We would like to highlight that this is preliminary research and the first of its kind to address naturalistic output with detailed discourse analysis which generated a very large body of data for each participant. For each participant a very large corpus of data is acquired, rather than dealing with single, disparate data points. This allows the examination of complex functions. Future research with more evenly distributed groups might be useful to confirm the veracity of these findings. Additionally, accounting for language dominance, atypical activation, and reorganisation is an important future direction. Procedures for lateralisation of language are typically only conducted as part of surgical decision-making, and as such this information was not uniformly available for this sample. Lastly, most assessments were completed via telehealth given COVID-19 lockdown conditions, where fewer rapport-building opportunities and technical disruptions [45] might promote brevity. Individual differences in responses to this format might have impacted their output and ultimately conflated findings, although these conditions were comparable among both groups.

## 5. Conclusions

These distinct patterns of impairment reflect a dynamic linguistic system taking shape under specific contextual conditions. In this functional system, micro- and macrolinguistic discourse components are differentially affected depending on task demands. Discourse in unconstrained conditions aligns closely with the clinician’s anecdotal reporting of verbosity in TLE which often does not map onto standardised assessment. This speaks to the demands of naturalistic clinical contexts where patients are asked open-ended questions. While this study has considered contextual demands on discourse, further research is essential to fully characterise the nuances of the neurolinguistic impairments in TLE beyond the single-word level. A future focus on circumstantiality in other constrained and unconstrained contexts is a viable avenue. Understanding language in naturalistic contexts provides better opportunities to consider how disturbances to language in TLE impact daily communication demands, and ultimately social and occupational functioning.

## Data Availability

All data produced in the present study are available upon reasonable request to the authors

## Data Availability

All data produced in the present study are available upon reasonable request to the authors

## Data Availability

All data produced in the present study are available upon reasonable request to the authors

## Acknowledgement of funding

This work was financially supported by the Australian Government Research Training Program Scholarship (Stipend and Fee offset) awarded by the Australian Commonwealth Government and the University of Melbourne to the first author.

## Author Contributions

**Fiore D’Aprano:** Conceptualisation (equal); Methodology (equal); Investigation (lead); Data Curation (lead); Formal Analysis (lead); Project Administration (lead); Visualisation (lead); Writing – Original Draft Preparation (lead); Writing – Review & Editing (equal).

**Charles B. Malpas:** Conceptualisation (equal); Methodology (equal); Formal Analysis (supporting); Resources (equal); Writing – Review & Editing (equal), Supervision (equal)

**Stefanie E. Roberts:** Data curation (supporting); Methodology (equal); Validation (lead)

**Michael M. Saling:** Conceptualisation (equal); Methodology (equal); Resources (equal); Writing – Review & Editing (equal), Supervision (equal)

**Disclosure of Conflicts of Interest:** None of the authors has any conflict of interest to disclose.

