## Supplementary material for "Macrolinguistic function in temporal lobe epilepsy: a reinterpretation of circumstantiality": Table S2

*Sample characteristics for neuropsychological variables*

| | Control (*n* =14) | | TLE (*n* =16) | | *p* | *r* |
|---|---|---|---|---|---|---|
| Variable | Median *(Q1,Q3)* | Range | Median *(Q1,Q3)* | Range | | |
| TOPF[a] | 0.75 (-0.02, 1.33) | 2.25 | 0.25 (-0.67, 0.67) | 3.00 | 0.06 | 0.42 |
| WASI-II $_{\text{Matrix Reasoning}}$ | 0.40 (0.00, 0.92) | 6.50 | 0.13 (-0.33, 0.58) | 4.50 | 0.47 | 0.16 |
| WASI-II $_{\text{Vocabulary}}$ | 1.00 (-0.18, 1.25) | 3.50 | 0.55 (0.00, 0.67) | 3.67 | 0.18 | 0.30 |
| WASI-II $_{\text{FSIQ}}$ | 0.61 (-0.25, 0.75) | 3.17 | 0.25 (-0.37, 0.55) | 4.50 | 0.27 | 0.24 |
| WAIS-IV Digits Forward[a] | 0.34 (-0.67, 0.92) | 3.67 | -0.33 (-0.33, 0.42) | 2.66 | 0.63 | 0.11 |
| WAIS-IV Digits Backward[a] | 0.00 (-0.67, 0.33) | 3.34 | 0.00 (-0.33, 0.67) | 2.66 | 0.85 | 0.04 |
| WMS-IV Logical Memory I[a] | 0.67 (-0.16, 1.33) | 4.00 | 0.33 (-0.33, 1.00) | 2.33 | 0.50 | 0.15 |
| WMS-IV Logical Memory II[a] | 1.00 (0.00, 1.00) | 3.66 | 0.33 (-0.42, 1.00) | 2.67 | 0.40 | 0.18 |
| RAVLT $_{\text{Total}}$[a] | 0.59 (-0.09, 1.52) | 3.57 | 0.11 (-0.23, 0.90) | 3.01 | 0.48 | 0.16 |
| RAVLT $_{\text{Post-Interference Recall}}$[a] | 0.22 (-0.58, 1.10) | 3.01 | 0.07 (-0.95, 0.78) | 3.76 | 0.47 | 0.16 |
| RAVLT $_{\text{Delayed Recall}}$[a] | 0.28 (-0.46, 1.26) | 4.34 | 0.30 (-0.90, 0.52) | 3.91 | 0.49 | 0.15 |
| WMS-R Easy^ | 12.00 (11.00, 12.00) | 3.00 | 11.00 (10.00, 11.25) | 5.00 | 0.12 | 0.33 |
| WMS-R Hard^ | 8.50 (6.25, 10.00) | 10.00 | 6.50 (6.00, 8.25) | 9.00 | 0.17 | 0.29 |
| WMS-R Easy Delay^ | 4.00 (4.00, 4.00) | 0.00 | 4.00 (4.00, 4.00) | 1.00 | 0.39 | 0.06 |
| WMS-R Hard Delay^ | 4.00 (3.25, 4.00) | 4.00 | 4.00 (3.00, 4.00) | 2.00 | 0.48 | 0.13 |
| Victoria Stroop $_{\text{Dots}}$[a] | -0.83 (-1.25, -0.33) | 2.33 | -1.17 (-1.33, -1.00) | 3.34 | 0.15 | 0.31 |
| Victoria Stroop $_{\text{Words}}$[a] | -0.50 (-0.92, -0.33) | 1.67 | -0.83 (-1.33, -0.33) | 2.33 | 0.24 | 0.25 |
| Victoria Stroop $_{\text{Colour}}$[a] | 0.00 (-0.92, 0.67) | 4.00 | -0.17 (-0.67, 0.33) | 2.00 | 0.54 | 0.13 |
| Victoria Stoop $_{\text{Interference}}$[a] | 0.83 (-0.83, 1.00) | 3.33 | 0.83 (0.33, 1.33) | 2.00 | 0.49 | 0.15 |
| BNT $_{\text{Total}}$[a] | -0.05 (-0.67, 0.55) | 3.03 | -0.77 (-1.54, 0.09) | 3.84 | 0.08 | 0.38 |
| BNT $_{\text{TOT States}}$^[a] | 17.00 (13.75, 23.50) | 22.00 | 24.00 (21.00, 27.00) | 17.00 | 0.02* | 0.48 |
| BNT $_{\text{Proportion TOT}}$^[a] | 0.28 (0.23, 0.40) | 0.37 | 0.40 (0.35, 0.45) | 0.28 | 0.02* | 0.48 |
| OLR (COWAT)[a] | -0.53 (-0.98, -0.04) | 4.01 | -1.15 (-1.58, 0.10) | 4.29 | 0.28 | 0.24 |
| Animals[a] | 0.67 (-0.02, 1.63) | 3.34 | -0.39 (-0.73, 0.25) | 2.79 | 0.006* | 0.59 |
| ANT $_{\text{Total}}$^ | 48.00 (48.00, 49.00) | 5.00 | 47.00 (45.00, 48.50) | 9.00 | 0.11 | 0.35 |
| ANT $_{\text{Latency}}$^ | 64.00 (59.50, 69.00) | 23.00 | 86.50 (70.75, 96.25) | 161.00 | 0.009** | 0.56 |
| ANT $_{\text{TOT States}}$^[a] | 11.50 (9.00, 14.50) | 18.00 | 22.00 (15.00, 27.25) | 41.00 | 0.02* | 0.50 |
| ANT $_{\text{Proportion TOT}}$^[a] | 0.23 (0.18, 0.29) | 0.36 | 0.44 (0.30, 0.55) | 0.82 | 0.02* | 0.50 |
| WFD Frequency Rating^[a] | 2.50 (1.25, 3.00) | 3.00 | 3.00 (2.25, 4.00) | 5.00 | 0.06* | 0.40 |
| WFD Distress Rating^ | 1.00 (1.00, 1.75) | 3.00 | 4.00 (3.00, 5.75) | 6.00 | 0.002** | 0.67 |
| VGT $_{\text{Correct Dominant}}$^ | 36.50 (34.25, 42.75) | 20.00 | 36.50 (32.00, 38.25) | 22.00 | 0.38 | 0.19 |
| VGT $_{\text{Correct Other}}$^ | 18.00 (15.00, 21.00) | 15.00 | 19.00 (16.00, 21.25) | 14.00 | 0.62 | 0.11 |
| VGT $_{\text{Latency}}$^ | 184.50 (164.25, 196.75) | 125.00 | 219.00 (183.75, 288.50) | 323.00 | 0.03* | 0.48 |
| HADS $_{\text{Anxiety}}$^[a] | 7.00 (5.25, 8.50) | 10.00 | 7.50 (6.75, 9.50) | 12.00 | 0.42 | 0.17 |
| HADS $_{\text{Depression}}$^ | 2.00 (1.00, 3.00) | 9.00 | 4.00 (4.00, 6.50) | 16 | 0.01* | 0.53 |

*Note.* TLE = Temporal Lobe Epilepsy; TOPF = Test of Premorbid Functioning; WASI-II = Wechsler Abbreviated Scale of Intelligence, Second Edition; FSIQ = Full Scale Intelligence Quotient; WAIS-IV = Wechsler Adult Intelligence Scale, Fourth Edition; WMS-IV = Wechsler Memory Scale, Fourth Edition; RAVLT = Rey Auditory Verbal Learning Test; WMS-R = Wechsler Memory Scale–Revised; BNT = Boston Naming Test; TOT = Tip-of-the-tongue; OLR (COWAT) = Orthographic Lexical Retrieval (Controlled Oral Word Association Test); WFD = Word Finding Difficulty; VGT = Verb Generation Task; ANT = Auditory Naming Task; HADS = Hospital Anxiety and Depression Scale. Represented as z-scores where normative data was available, raw data are indicated by^. Latencies are expressed in seconds. Group differences computed using Mann-Whitney U test, effect sizes *r*, where * = p <.05, ** = p <.01. . [a]Suggests that data does not violate assumptions of normality on Shapiro-Wilk.
