## Supplementary material for "Macrolinguistic function in temporal lobe epilepsy: a reinterpretation of circumstantiality": Table S3

*Discourse features of interest and their coding criteria*

| Discourse Feature | Description |
|---|---|
| *"and"* | Any instance of the word "and", the semantically least marked conjunction in place of other conjunctions. To be counted separately and then included in count of total connectives. |
| *Abandonment* | Discarding all of the interrupted utterance and replacing it entirely |
| *Attempted/incomplete ties* | The number of cohesive ties which are either ambiguous or incorrect (e.g. number, gender, meaning) are tallied and the ratio to the total number of cohesive ties is calculated. |
| *Clarity Disruptors* | Composite metric includes deictic terms, circumlocution, empty phrases, and indefinite terms. Deictic terms are a word or phrase (e.g., this, that, these, those, now, then, here) that points to the time, place, or situation in which a speaker is speaking. Empty phrases are common idioms or idiosyncratic fillers not contributing any content to the discourse (e.g. "and so on and so forth", "you know", "like", "but yeah"). Indefinite terms are highly non-specific nouns or terms, (e.g. "thing", "something", "stuff") |
| *Clausal embedding* | Count of number of embedded clauses, as either left- or right-branching. In left-branching clauses, the embedded/dependent clause interrupts the main clause (comes first), e.g., "When I saw the cars on the road, I was really frightened." In right-branching structures, the main clause is followed by the embedded clause, e.g., "I was really frightened when I saw the cars on the road" |
| *Cohesion Disruptors* | A composite metric including count of 'ands', other referents (missing or attempted ties), referent repetition errors - being those where a referent or its pronoun is used inconsistently for the same object or character. |
| *Conjunctions* | A cohesive relation occurring between clauses and specifies the way in which what is to follow is systematically connected to what has gone before, e.g. but, or, so, because. (excluding and) |
| *Connectives* | Incorporating instances of conjunctions and "ands", thereby representing all sentence connections |
| *Cookie core units* | Total number of novel core content units as per Yorkston and Beukelman (1977) |
| *Deictic terms* | a word or phrase (such as this, that, these, those, now, then, here) that points to the time, place, or situation in which a speaker is speaking (excludes the use of "that" as a relative pronoun). |
| *Empty phrases* | Common idioms or idiosyncratic fillers not contributing any content to the discourse (e.g., "and so on and so forth", "you know", "like") |
| *False start* | A phrase or sentence which is revised to modify, correct or clarify what the speaker has already said and may include an editing expression such as "I mean", etc. E.g "I went to town on Thursday I mean on Friday". Includes abandonments, incomplete mazes, word-choice corrections, part-word productions, and repetitions. |
| *Filler* | Any aspect of speech which is non-communicative and serves no addition to content (e.g., "um", "ah", "er") |
| *Fluency disruptors* | Composite metric of clarity disruptors, false starts and repairs (including abandonment, incomplete mazes, part-word production, repetitions), hesitations (pauses and fillers) and word finding difficulties |
| *Incomplete mazes* | A phrase or sentence which is begun but abandoned, leaving the thought incomplete and signifying a shift in thought, e.g. "After going home.. well first I went shopping" |
| *Indefinite words/terms* | Highly non-specific nouns (e.g., "thing", "something", "stuff") |
| *Non-progression error* | Instances where information that has already been presented is repeated or redundant, or where content is irrelevant to the task goals |

| | |
|---|---|
| *Other referents* | A composite metric including attempted/incomplete ties and missing referents - being those where the referent or subject is absent or omitted from the statement. |
| *Part word production* | The production or repetition of syllables and sounds within a word, e.g. "He went ho- home first" |
| *Pause (unfilled)* | Any duration of silence (>250ms) |
| *Pause duration* | Collective length of time spent pausing |
| *Postponement* | Interrupting an utterance so additional material may be inserted, with a return to the original utterance later on, e.g. "and they thought- there was pig's blood in it and they thought that there was somebody hurt. But it was the pig". |
| *Production rate* | Words spoken per second |
| *Reference correction* | Replacing a noun or pronoun so as to specify a referent more exactly, for the purpose of disambiguation, e.g. "Gerald and Kevin threw a snowball at me and I got him right in the face- Kevin right in the face", or where a pronominal reference may be replaced by a noun phrase, e.g. "we went to- uh me and Jess went to Aunt Carol's" |
| *Referential ties* | Instances where an individual refers to another object/person using pronouns. Code anaphoric and cataphoric referential ties separately. Anaphoric = noun usage precedes pronoun introduction. Cataphoric = pronoun usage precedes introduction of noun. |
| *Repetition (single and multi-word)* | Instances of repetition of a previous word or words of the utterance before carrying on, e.g. "I have- I have new shoes on". Single vs multi-word to be coded separately, as well as utterance-initial and mid-utterance occurrences. |
| *Repetition errors* | Errors associated with the semantic relatedness of different parts of a narrative, and are marked by the inconsistent used of pronouns, or referents for the same object or character. These correspond with instances of inconsistent referential ties. |
| *Sample length* | Count of words spoken for a given task or trial. Includes all completed words uttered (excluding part words and fillers, although including repetitions) |
| *Spontaneous duration* | Length of time producing output for a given task or trial |
| *Syntactic complexity* | Composite metric: clausal embedding, postponement, conjunctions, syntactic corrections |
| *Syntactic correction* | Refers to instances of replacing a content word with the same word of a different morphological form, rearranging word order, inserting appropriate function words, altering noun-pronoun agreement, or any other repair that improves the syntactic quality of the utterance. E.g. "He felt like there's bells in his ears and he can't- couldn't hear". |
| *Syntactic simplicity* | Composite metric: missing referents, 'ands', and syntax errors |
| *Syntax error* | Count of errors to syntax including incomplete utterances, missing definite article, subject, or referent |
| *Total statements* | Number of statements, where single statement refers to a predicate and its corresponding arguments |
| *Word choice correction* | Replacing a content word with another of the same morphological form (e.g., "yesterday I leant- borrowed my friend's car") |
| *Word finding difficulties* | Instances where an individual is unable to produce the correct word, and either uses circumlocutory language or non-specific language to attempt to describe it |
