## Supplementary material for "Macrolinguistic function in temporal lobe epilepsy: a reinterpretation of circumstantiality": Table S4

*Description and example of TLE output in constrained output (Cookie Theft)*

| *Cookie Theft measure* | *TLE impairment* | *TLE example output* |
|---|---|---|
| Pause duration | Spend a longer duration pausing | |
| Production rate (words/second) | Produce fewer words per second. Where the production rate is slower, language output is considered to be less fluent. | |
| Fluency disruptors | Produce more disruptions of fluency than healthy controls. These include empty phrases, indefinite terms, deictic terms, false starts and repairs | "standard kitchen benches and all of that" (empty phrase)<br><br>"fall and break his neck (0.60) but anyway (0.31) that's just the way it is"(empty phrase)<br><br>"trying to get a cookie as well while the boy has (0.26) been going on (0.26) this (0.87) chair"(deictic term)<br><br>he's going to fall off that chair there (deictic term)<br><br>"um (1.60) couple of- couple of things sitting out on the counter (0.25) to be washed" (indefinite term) |
| Pauses (non-grammatical) | Produce more instances of pauses in non-grammatical positions, i.e., occurring in the middle of a clause rather than at a natural grammatical juncture. This creates disruption in the fluency of output | "and then we've got (0.62) a little girl"<br><br>"and his (0.25) sister or a little girl is (0.39) um (0.60) putting her (1.12) yeah putting her hand up" |
| Disrupted Cohesion | Have significantly more elements that disrupt the overall cohesion of their language output, i.e., there are more disruptions to interconnectedness between sentences. This includes the use of other referents that are ambiguous, incomplete, or missing and errors in the consistency of referents. Whereas controls tend to avoid the use of referent repetition errors, by providing consistent referents where necessary and otherwise using anaphoric referents such as "he" and "she" to complete referential ties. | "um just having a really good talk" (missing referent)<br><br>"a brother and sister are trying to eat cookies but maybe the sister's like I'm gonna- (1.05) I'm gonna (0.25) tell (0.27) her if you don't give me one" (ambiguous referent)<br><br>"boy" "young boy" "fella" used interchangeably in same elicitation instead of single referent and subsequent ties such as "he"(referent repetition error) |
