## Supplementary material for "Macrolinguistic function in temporal lobe epilepsy: a reinterpretation of circumstantiality": Table S5

*Description and example of TLE output in unconstrained output (Typical Day)*

| *Measure Typical Day* | *TLE impairment* | *TLE example output* |
|---|---|---|
| Sample length (total words) | Produce output containing more words overall | |
| Spontaneous duration (seconds) | Produce output that has a longer duration overall | |
| Pause duration | Spend a longer duration pausing | |
| Duration excluding pauses | Even when excluding the amount of time spent pausing, those with TLE produce a longer duration of output | |
| Pauses (non-grammatical) | Produce more instances of pauses in non-grammatical positions, i.e., occurring in the middle of a clause rather than at a natural grammatical juncture. This creates disruption in the fluency of output | "and um I uh (0.49) make myself (0.25) um some breakfast"<br><br>"so I'm expecting a call from (0.25) mechanics to say (0.47) uh this car needs that done this car needs that done" |
| Proportion Non-Progression to Task-On Novel Units (non-progression includes task-off novel, task-off repeated, and task-on repeated-redundant units) | Produce a significantly higher proportion of statements that do not progress content relative to those that are novel and related to the task. As such, those with TLE produce more statements that are vague, repetitive, or unrelated to the task at hand. The output is less informative overall | "um (0.66) a typical day in my life is that I wake up at around about seven thirty (0.31) and I feed my dog (0.89) and uh (0.25) he's uh champing at the bit to get fed 'coz he only gets fed once a day (task-off novel) (0.41) so I feed him"(task-on repeated-redundant) |
